# Neurodoron® in patients with neurasthenia – A randomized, double-blind, placebo-controlled clinical trial

**DOI:** 10.1101/2022.09.02.22279531

**Authors:** Juliane Hellhammer, Luitgard Spitznagel-Schminke, Cristina Semaca, Rebecca Hufnagel

**Affiliations:** Contract Research Institute daacro, Science Park, Max-Planck-Strasse 22, 54296, Trier, Germany; Weleda AG, Möhlerstrasse 3-5, 73525 Schwäbisch Gmünd, Germany

**Keywords:** Neurodoron®, anthroposophic medicine, nervous exhaustion, neurasthenia, stress, irritability, nervousness, randomized controlled clinical trial

## Abstract

1

**Introduction:** The term neurasthenia has been introduced in the late 19th century. Stress was indicated as one of the main triggers. Many treatment options are available to reduce the associated symptoms. Complementary and alternative medicine (CAM) is widely used due to its long tradition and positive safety profile. This Phase IV placebo-controlled clinical trial was designed to demonstrate efficacy and safety of the CAM-product Neurodoron® in patients with neurasthenia.

**Methods:** This monocentre, randomized, double-blind, placebo-controlled, parallel-group clinical trial was conducted in a dedicated outpatient German trial site. Women and men aged 18 and above were randomized to receive either Neurodoron® or matching placebo if they reported typical symptoms of neurasthenia and a severe psychiatric disorder could be excluded. The primary objectives were a reduction in characteristic symptoms of nervous exhaustion and perceived stress as well as improvement in general health status after 6 weeks of treatment.

**Results:** 204 patients underwent screening, 78 were randomized in each treatment group, and 77 patients each received treatment (intention-to-treat (ITT) population = 154 patients). For none of the primary efficacy variables, an advantage in favor of Neurodoron® could be demonstrated in the pre-specified analysis (p-values between 0.505 to 0.773, Student’s t-test). In a post-hoc analysis of intra-individual differences after 6 weeks treatment, a significant advantage of Neurodoron® vs. placebo was shown for characteristic symptoms of nervous exhaustion (irritability (p = 0.020); nervousness (p = 0.045), Student’s t-test). Adverse Event (AE) rates were similar between treatment groups, in both groups 6 AEs were assessed as causally related to treatment (severity mild or moderate). No AE resulted in discontinuation of treatment.

**Conclusion:** A significant improvement of neurasthenia was seen for the total study population at the end of the treatment period. Superiority of Neurodoron® vs. placebo could not be demonstrated with the pre-specified analysis. However, the post-hoc analysis suggests Neurodoron® as a beneficial option over placebo for the treatment of neurasthenia, especially given its confirmed markedly good safety.

## 2 Introduction

Originally, “neurasthenia”, meaning exhaustion of the “nerve strength”, was introduced – presumably independently – by two US-American physicians, Edwin Holmes Van Deusen and George Miller Beard, at the end of the 19^th^ century [1, 2]. Nowadays, the term neurasthenia is more closely associated with the New York neurologist Beard, who primarily saw the city’s upper middle-class patients [3]. He used the term neurasthenia to refer to an acute or mainly chronic, functional condition of the nervous system, characterized by a range of predominantly somatic symptoms, including headaches, dyspepsia, lack of concentration, irritability, insomnia, and hopelessness in the absence of any organic disease [1, 4]. For Beard, stress in private or business life due to modern civilization with a shift from manual labor to mental work was one of the main triggers for neurasthenia [5]. This cause, combined with the scientific approach of attributing neurasthenia to a primarily physical disease rather than a – at these times – stigmatizing psychic illness, resulted in a well-accepted disease that could be treated by a neurologist (and not by a psychiatrist). A vast group of nervous disorders was initially assigned to neurasthenia, due to its vague and broad definition and its social acceptance [6, 7]. But over the further decades its importance diminished, partly also due to this broad definition, but also because Beard’s theory of “nerve strength” could not be proven on a physiological level [6].

However, neurasthenia experienced a revival with a shift to mental symptoms [8]. In 1977, the diagnosis neurasthenia was included in the manual of the international statistical classification of diseases, injuries, and causes of death [9]. Nowadays, the term neurasthenia is used for patients with chronic exhaustion or fatigue who have characteristic symptoms and in whom organic causes can be excluded [4, 10, 11].

Data on frequency of neurasthenia vary: an international WHO-multicenter study gives numbers between 1.3% and 10.8% in the general population [12]. In a WHO-report on health care, the prevalence of pure neurasthenia ranged between 0.3% (Athens and Bangalore) and 3.7% (Manchester). A long-term study in Switzerland suggests that prevalence increases with age (0.7% for 22-year-olds to 3.8% for 41-year-olds) [13]. The persistent exhaustion often leads to absenteeism and thus causes economic costs [14].

Treatment options for chronic fatigue include cognitive and behavioral therapies, as well as exercise therapy or a gradual increase of activity [15]. Henningsen, Zipfel, and Herzog propose a graded therapeutic approach including the administration of pharmacotherapy as a symptomatic treatment [16] but currently there is no internationally accepted standard therapy for neurasthenia.

There is minor evidence for the efficacy of antidepressant and immunoglobulin therapies, no evidence could be found for an efficacy of corticosteroid and interferon therapies [15, 16]. Some of the benzodiazepines have shown efficacy in establishing sleep maintenance, but have side effects, e.g. next-day sedation, and can only be used to a limited extent or for a limited period of time due to the addictive and habituation effect [17]. Even in the absence of randomized controlled clinical trials (RCTs), resting, sparing behavior, and inactivity are considered clearly contraindicated [15].

However, drug treatment to alleviate the symptoms is feasible. The characteristic symptoms, such as nervousness, irritability, restlessness, and sleep disorders, can be well influenced with complementary and alternative medicine (CAM) approaches, such as natural compounds, a safe alternative to conventional medicine [18]. The effect is shown by various clinical studies for different herbal and homeopathic remedies [19-24]. However, no data from RCTs had been available yet.

The present drug, Neurodoron, is a CAM representative from anthroposophic medicine. Data from a non-interventional study (NIS) performed with physicians in Germany from 2008 to 2009 confirmed that it is a valuable alternative for treatment of stress-related nervous fatigue [24]. Based on these findings the objectives of the presented clinical trial were to demonstrate efficacy and safety of Neurodoron® by means of an RCT, a method regarded as gold standard in clinical research. Following the RCT, a pharmacy-based NIS was conducted between 2014 and 2016, which confirmed the beneficial effects of Neurodoron® in self-medication [25].

Neurodoron^®^ is an anthroposophic medicinal product that is supposed to support the body systems in order to rebalance mind and body. Anthroposophic medicine, as part of CAM, is an integrative multimodal treatment system based on a holistic understanding of disease and treatment, which was developed in the early 1920s in Germany and Switzerland [26, 27]. Meanwhile, a curriculum for becoming a certified anthroposophic physician is available in numerous countries. Europe is most strongly represented, but also countries from all other continents, except for Africa and the Antarctic. The training takes different lengths of time, e.g. 3 years in Germany, and a conventional medical degree is a prerequisite. Anthroposophic medicines are also prescribed by conventional physicians without special training [27].

In the early 1950s, Dr. Kurt Magerstaedt developed Neurodoron® to support people to better cope with exam stress. In 1954, the product was first launched in Germany, followed by Switzerland, Austria, France, Ukraine, Italy, and UK. Since 2007, Neurodoron® has been listed in Germany with the indication nervous exhaustion and metabolic weakness, based on case reports from medical practitioners indicating that Neurodoron® seems to be efficient in treating stress related health complaints [28]. Neurodoron® contains *Aurum metallicum praeparatum, Kalium phosphoricum*, and *Ferrum-Quarz* (Weleda AG, Schwäbisch Gmünd, Germany). According to the anthroposophical approach, the three components of Neurodoron® are to support the three existing systems, namely the nervous-sensory system, the rhythmic system and the metabolic-limb system, thus helping to rebalance mind and body.

## 3 Methods

### 3.1 Clinical trial design

This clinical trial was a monocentric, randomized, double-blind, placebo-controlled parallel group phase IV superiority trial, conducted at a dedicated clinical trial site in Berlin, Germany. The clinical trial was coordinated by the contract research organization ACM Allied Clinical Management GmbH. Prior to enrollment of the patients, the trial was approved by the relevant competent authorities and the ethics.

Four trial visits and a 6-weeks treatment phase were scheduled. Written informed consent was obtained from each patient at the screening visit, prior to trial enrollment. If a patient was eligible to participate in the trial, randomization and initiation of treatment occurred at baseline (Visit 1). At baseline, and at the two consecutive trial visits – after 2 (Visit 2) and another 4 weeks (Visit 3 / final visit) – patients rated 12 characteristic symptoms, their perceived stress (Perceived Stress Questionnaire, PSQ) and their general health status (Short Form Health Survey, SF-36). Blood samples for safety laboratory were taken at screening and final visit. All procedures per trial visits and the flow chart are shown in the supplemental material I.

### 3.2 Clinical trial population

Main eligibility criteria were based on the diagnosis F48.0 (ICD-10 [29]), i.e. persistent and distressing complaints of feelings of exhaustion after minor mental effort or persistent and distressing complaints of feelings of fatigue and bodily weakness after minor physical effort for at least 3 months. Furthermore, at least one of the following characteristic symptoms had to be present: muscular aches and pains, dizziness, tension headaches, sleep disturbance, inability to relax and irritability. Male and female patients aged at least 18 years were excluded if they reported an organic cause for the complaints or presented with a certain severe psychiatric disorder. A full list of inclusion and exclusion criteria is available in the supplemental material II.

### 3.3 Randomization and intervention

Trial patients were randomly assigned to either Neurodoron® or placebo in a 1:1 ratio. The randomization list was generated using a validated computerized system (EDP random number generator (SAS Version 9.2; RANUNI function)) by an independent institute. The block size was not revealed to the investigator. Investigational medicinal product (IMP) containers were numbered sequentially. Each eligible patient was assigned to the next available IMP container number by the investigator at the investigational site. Neither investigator nor patient could identify group assignment based on the IMP container. The randomization list was broken only after finalization of the statistical analysis plan and the subsequent database lock.

### 3.4 Investigational medicinal product (IMP)

On the market for more than 50 years and known as *Kalium phosphoricum comp*. (Weleda AG, Schwäbisch Gmünd, Germany) until 2007, the IMP has been listed under the name Neurodoron® as an anthroposophic medicine with the indication of nervous exhaustion and metabolic weakness (German registration number 6646311.00.00, German Commission C monograph [30]). One tablet of Neurodoron® contains the following active ingredients: 83.3 mg *Aurum metallicum praeparatum trituration (trit*.) D10, 83.3 mg *Kalium phosphoricum trit*. D6, 8.3 mg *Ferrum-Quarz trit*. D2 (excipients: wheat starch and lactose). The corresponding matching placebo was identical in form, color, size, taste and smell and contained the same ingredients, except for the active ingredients. Tablets were to be taken orally 3 times a day with a little liquid for 6 weeks.

### 3.5 Objectives

The aim of this trial was to strengthen the evidence for the safety and efficacy of Neurodoron® on nervous exhaustion.

#### 3.5.1 Primary objectives

Primary objective of this superiority trial was to demonstrate efficacy of Neurodoron® by reducing characteristic symptoms of nervous exhaustion and perceived stress and by improving the general health status. Respective outcome measures (characteristic symptoms, perceived stress questionnaire (PSQ), SF-36) were assessed at trial end and compared to treatment start. Superiority of the verum was considered evident if a statistically significant improvement of Neurodoron® over placebo could be demonstrated, based on the mean sum score of the characteristic symptoms.

In a post-hoc analysis, also the single symptoms and the subscales of the PSQ and SF-36 were analyzed.

#### 3.5.2 Secondary objectives

Secondary objectives of this trial were to assess the efficacy of Neurodoron®:

- on intensity of tedium (Tedium Measure) at Visits 1, 2, and 3
- based on the efficacy ratings of both patients and physicians at treatment end.

In addition, safety of the Neurodoron® treatment was to be confirmed based on the type and frequency of clinically relevant laboratory value changes at treatment end vs. screening. Additionally, the frequency and severity of (serious) adverse events throughout the trial and the tolerability ratings of both patients and physicians at treatment end were assessed for safety evaluation.

### 3.6 Measurements of trial parameters

#### 3.6.1 Demographic and anthropometric data

Age, gender, height, weight, smoking status, marital status and professional situation were recorded.

#### 3.6.2 Characteristic symptoms related to nervous exhaustion

Characteristic symptoms of nervous exhaustion were rated on a four-level Likert scale ranging from “0 = absent” to “3 = severe” and a symptom sum score was calculated (maximum value 36). 12 symptoms were assessed: irritability, restlessness, nervousness, listlessness, depressive mood, mood swings, anxiety states, troubles to concentrate/lack of concentration, headache, sleep disorders, digestive disorders, muscular pain/tensions.

#### 3.6.3 Perceived Stress Questionnaire (PSQ) [31]

For assessment of individually perceived stress, the revised German version of the PSQ-20 was used, a reliable and validated tool in stress research [32, 33]. It contains 20 items which can be assigned to four subscales (worries, tension, joy, and demands). The patients make their assessment on a four-level Likert scale ranging from “1 = almost never” to “4 = usually”. Scores for the subscales and the total score are obtained by calculating the mean and a subsequent linear transformation to values between 0 and 100. For calculation of the total score the values for “joy” are inverted. Thus, higher total scores indicate more severe perceived stress.

#### 3.6.4 Short Form-36 Health Survey SF-36 [34]

Health-related quality of life was evaluated using the validated SF-36. This questionnaire is composed of 36 items (with different scores from 1-2 to 1-6) and assesses mental, physical and social health aspects. The 36 items of the SF-36 are summarized in eight subscales (vitality, physical functioning, bodily pain, general health perceptions, physical role functioning, emotional role functioning, social role functioning, and mental health). In addition, the subscales are computed into a physical and mental sum score using SAS and SPSS. For calculation of the scores, the recorded numeric values are linearly transformed to a 0 to 100 range (all items are converted in the same direction, so the higher the score the better the quality of life). In a second step, items in the same scale are averaged.

#### 3.6.5 Tedium Measure [35]

The Tedium or Burnout Measure was developed by Pines and Aronson [35] and is used for self-rating of burnout. This tool consists of 21 items that are scored on a 7-level Likert scale ranging from “never” to “always”, assessing the degree of physical, emotional and mental exhaustion. The Tedium Measure is calculated according to the authors’ specifications and can reach values between 2.1 and 5.9. High values of the Tedium Measure describe a high extent of tedium or burnout. Pines et al. [36] offer interpretations for the test scores: values ≤ 3 suggest no problems, values between 3 and 4 indicate a high risk, and values > 5 an acute crisis.

#### 3.6.6 Efficacy and tolerability rating by patient and physician

Trial patients and investigators rated the efficacy and tolerability of the IMP separately on a four-level Likert scale ranging from “very good” to “unsatisfactory”.

#### 3.6.7 Safety

The number and severity of adverse events after intake of IMP was recorded for both groups independently from the causality. The laboratory values at screening and treatment end were examined as an additional safety parameter, as was the tolerability ratings by the patient and physician (3.6.6), and physical examination (baseline and treatment end) and vital signs (all trial visits).

### 3.7 Sample size and statistical analysis

The calculation of the sample size was based on the characteristic symptoms associated with nervous exhaustion and resulted in 182 patients to be enrolled to obtain 146 evaluable patients, 73 in each treatment group (assumptions: 25% advantage of Neurodoron® over placebo at treatment end (with Neurodoron® (sum symptom score/mean): 7.1 (± 5.1), drop-out rate: 20% (α: 0.05, β: 0.2)).

All statistical analyses were performed with SAS Version 9.2 (SAS Institute Inc., Cary, NC, USA) or SPSS 20 (SPSS Inc., Chicago, IL, USA).

All demographic and baseline data were descriptively analyzed. The homogeneity of treatment groups was tested.

For confirmatory analysis, the three primary efficacy variables were tested in hierarchical order in the intention-to-treat (ITT) population, i.e., in all patients who had administered at least one dose of the IMP and had no missing values. Therefore, depending on the variable analyzed, the number of patients considered for analyses varied slightly (max. n = 154). All analyses were carried out using Student’s t-test for two independent samples on the same one-sided alpha-level of 0.025 (equivalent to 0.05 two-sided). All analyses performed for the primary endpoint were also performed with the per-protocol (PP) population (n = 131).

A post-hoc analysis was performed for the three primary efficacy variables with SPSS 22 and Statistica 14. While the original analysis was planned on the sum scores, the post-hoc analysis was performed for evaluation of intra-individual differences based on single symptoms (characteristic symptoms), and subscales (PSQ, SF-36), respectively (Student’s t-test for two independent samples). The two treatment groups were also analyzed over time using analysis of variance with repeated measures (repeated measures ANOVA) with three time points (Visit 1, Visit 2, Visit 3).

The secondary efficacy variables were exploratively analyzed. For this purpose, group comparisons were carried out using statistical tests (Tedium Measure: Student’s t-test for two independent samples, efficacy rating by patient and physician: chi-square test); the results of these analyses were indicative yet not confirmatory.

## 4 Results

Between September 2011 and July 2012, 204 patients were screened, 156 patients were invited for baseline/visit 1, 154 patients underwent treatment with n = 154 in the ITT and safety population (SP), and n = 131 in the PP population (fig. 1). The last trial visit took place in September 2012. In this trial, 154 patients were randomized and received allocated treatment of which 113 were female (73.4%) and 41 male (26.6%). Mean age of the patients was 53.1 (± 13.1). The mean treatment duration was 41.9 ± 4.5 days (Neurodoron® 41.7 ± 5.0 days, Placebo 42.1 ± 3.9 days), the median treatment duration was 42.0 days, range 10.0 – 50.0 days (Neurodoron® 42.0 days, range 10.0 – 48.0 days, Placebo 42.0 days, range 19.0 – 50.0 days). When comparing baseline characteristics between the Neurodoron® and the placebo group, patients of the placebo group were more often ex-smokers (32.5%) compared to patients of the Neurodoron® group (22.1%), but there were more smokers in the Neurodoron® group (26.0% vs. 15.6%). Patients of the Neurodoron® group were more often employed workers compared to patients of the placebo group (40.3% vs. 27.3%). The residual baseline characteristics did not differ 10% between both groups (tab. 1).

**Tab. 1:**
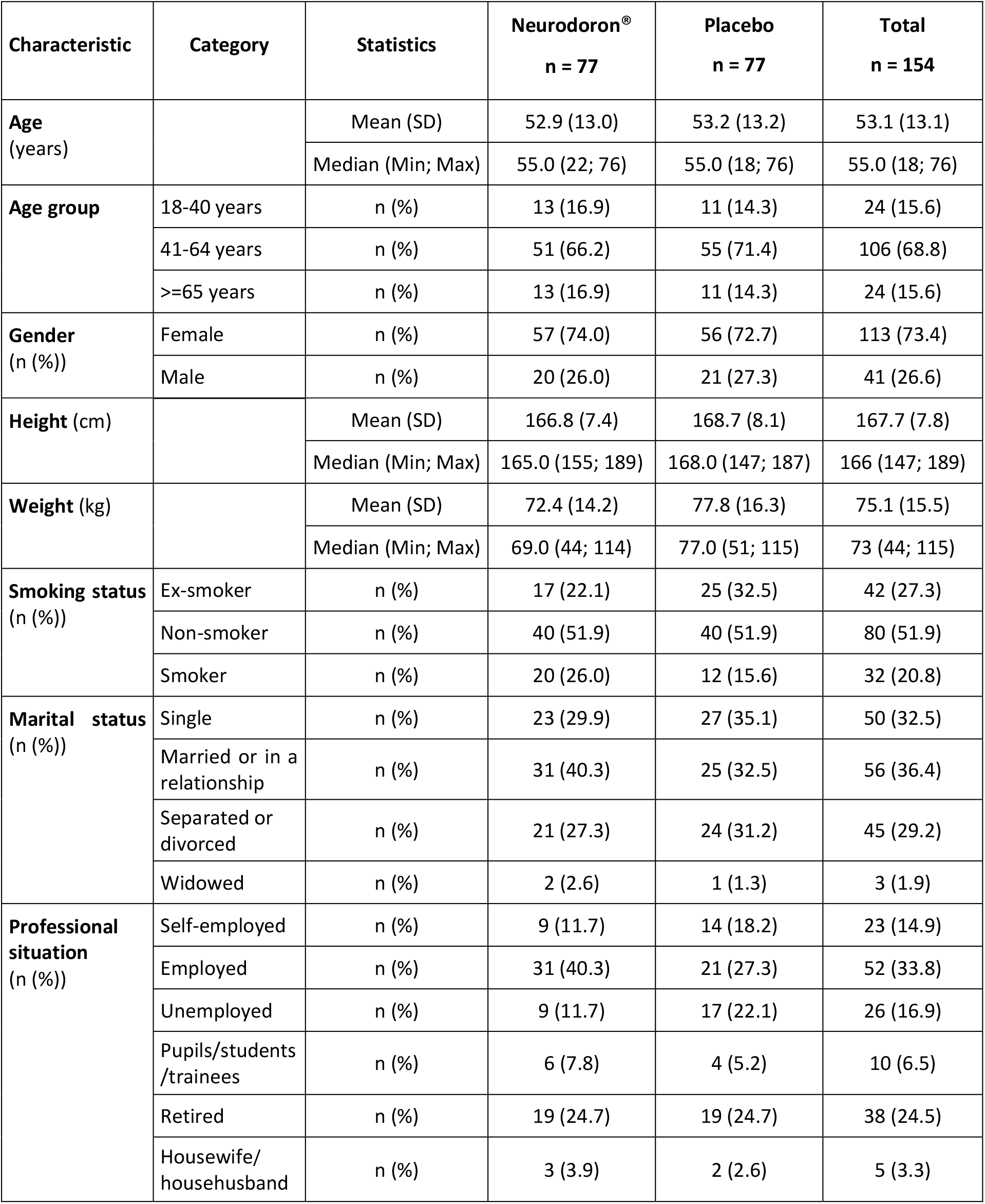
Characteristics of trial patients, ITT-population (n = 154)

**Fig. 1.**
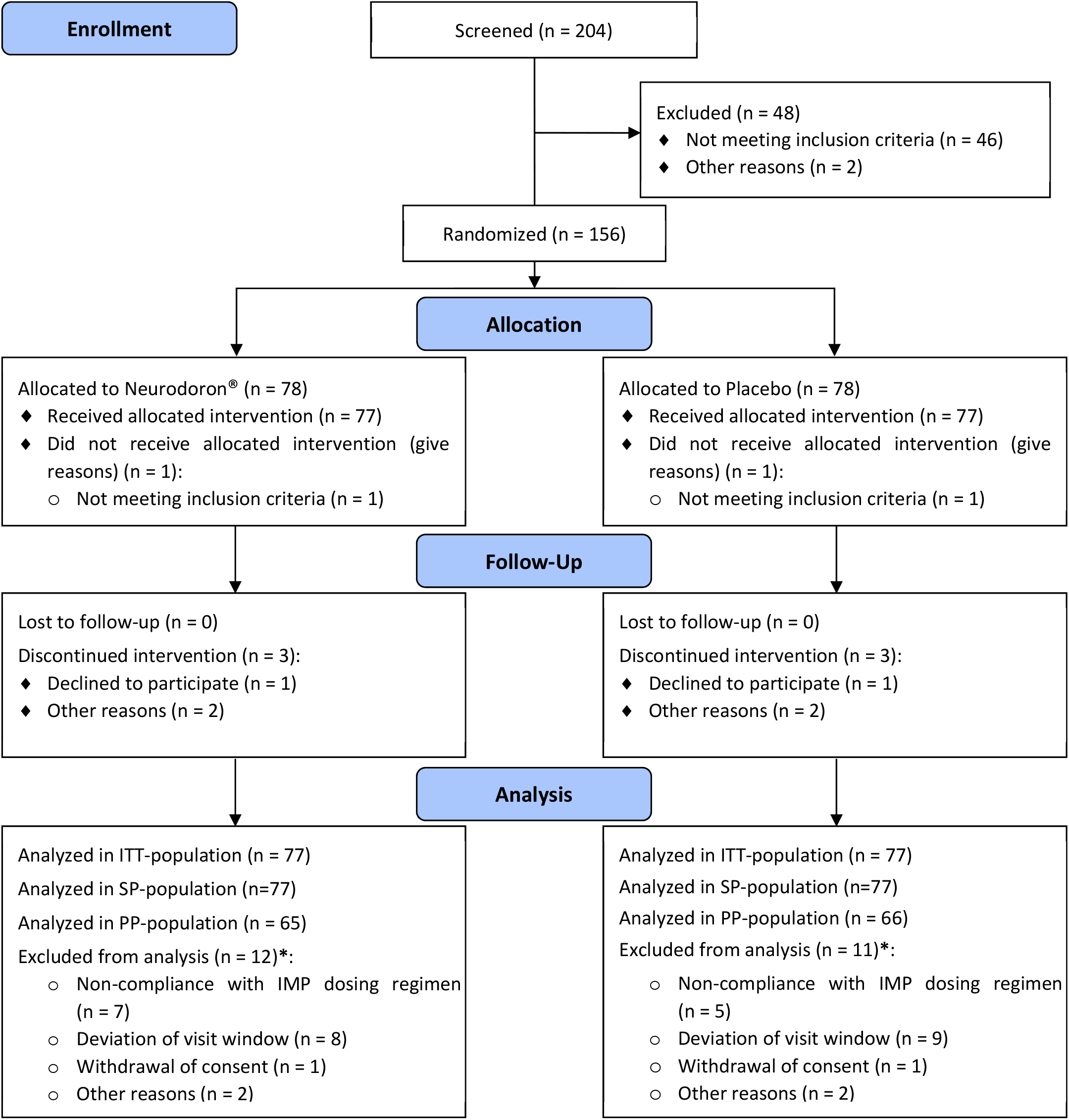
CONSORT Flowchart of the trial patients showing the distribution of the patients from initial assessment to analysis of trial data.

### 4.1 Reasons for neurasthenia

Reasons for neurasthenia were comparable between the treatment groups (Tab. 2). Patients (n = 154) reported work or school as the most common cause of neurasthenia (Neurodoron: 53.2%, Placebo: 49. 4%), followed by housekeeping (Neurodoron: 33.8%, Placebo: 31.2%). Apart from reasons associated with work/school or all reasons associated with the familial situation, other not further specified reasons were frequently documented (Neurodoron: 45.5%, Placebo: 37.7%).

**Tab. 2:** Reasons for neurasthenia, ITT-population (n = 154)

| Reason(s)* | Statistics | Neurodoron®<br>n = 77 | Placebo<br>n = 77 | Total<br>n = 154 |
| --- | --- | --- | --- | --- |
| Work or school | n (%) | 41 (53.2) | 38 (49.4) | 79 (51.3) |
| Housekeeping | n (%) | 26 (33.8) | 24 (31.2) | 50 (32.5) |
| Raising children | n (%) | 10 (13.0) | 14 (18.2) | 24 (15.6) |
| Care for relatives | n (%) | 4 (5.2) | 7 (9.1) | 11 (7.1) |
| Care for disabled | n (%) | 0 (0.0) | 1 (1.3) | 1 (0.6) |
| Partnership conflict / divorce | n (%) | 4 (5.2) | 9 (11.7) | 13 (8.4) |
| Other reasons | n (%) | 35 (45.5) | 29 (37.7) | 64 (41.6) |

\*multiple entries possible

### 4.2 Primary efficacy analysis

#### 4.2.1 Efficacy on 12 characteristic symptoms of nervous exhaustion

The mean symptom sum score at visit 3 did not differ significantly in the a priori analysis between the placebo and the Neurodoron® group (p = 0.728, fig. 2, tab. 3).

**Tab. 3:**
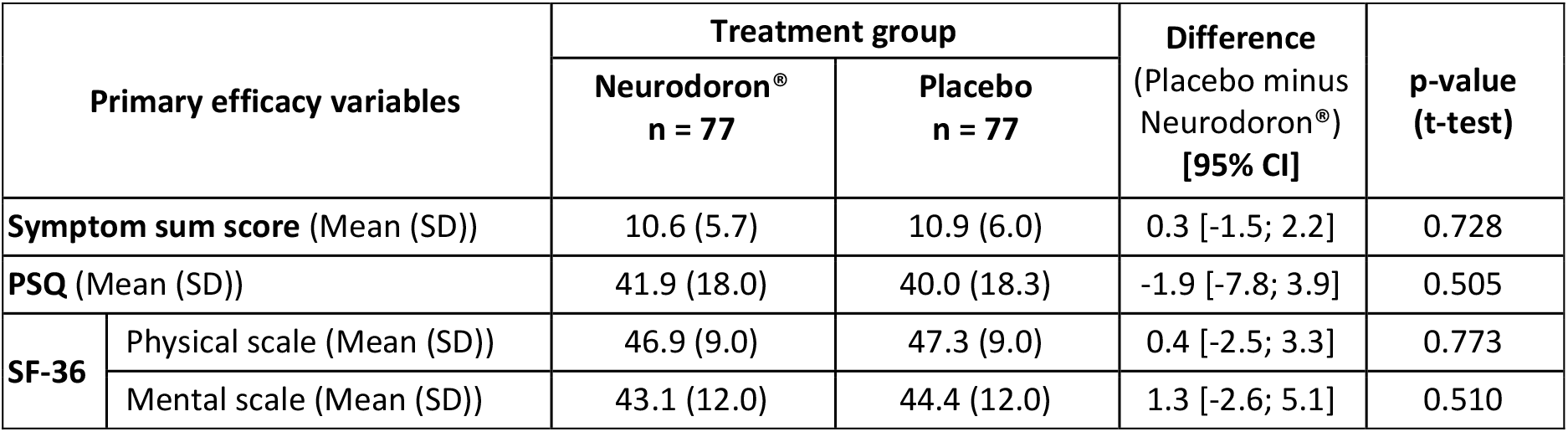
Primary efficacy results at visit 3 (after 6 weeks of treatment), ITT population (n = 154)

| Primary efficacy variables |  | Treatment group |  | Difference<br>(Placebo minus<br>Neurodoron®)<br>[95% CI] | p-value<br>(t-test) |
| --- | --- | --- | --- | --- | --- |
|  |  | Neurodoron®<br>n = 77 | Placebo<br>n = 77 |  |  |
| Symptom sum score (Mean (SD)) |  | 10.6 (5.7) | 10.9 (6.0) | 0.3 [-1.5; 2.2] | 0.728 |
| PSQ (Mean (SD)) |  | 41.9 (18.0) | 40.0 (18.3) | -1.9 [-7.8; 3.9] | 0.505 |
| SF-36 | Physical scale (Mean (SD)) | 46.9 (9.0) | 47.3 (9.0) | 0.4 [-2.5; 3.3] | 0.773 |
|  | Mental scale (Mean (SD)) | 43.1 (12.0) | 44.4 (12.0) | 1.3 [-2.6; 5.1] | 0.510 |

**Fig. 2.**
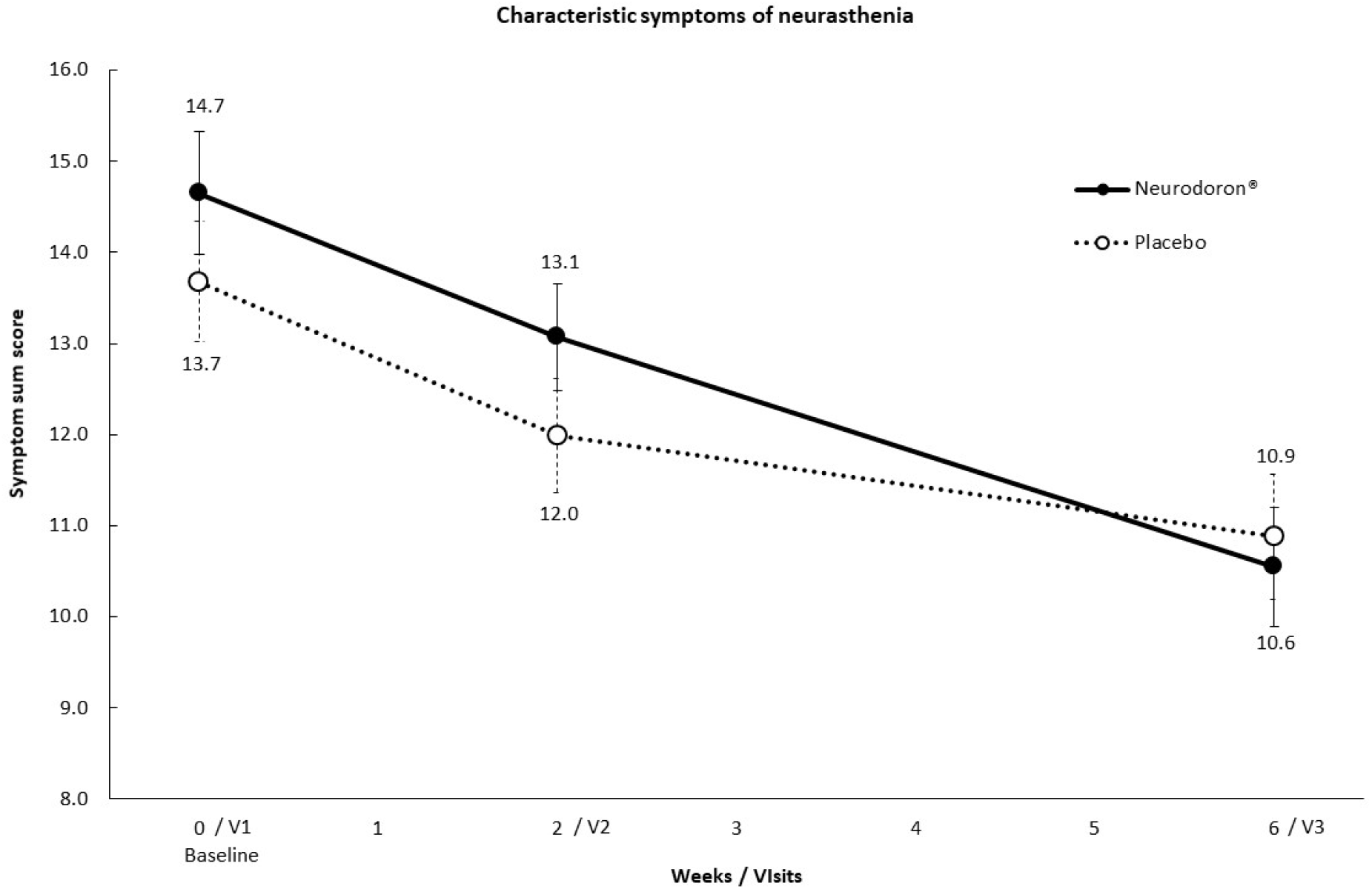
Symptom sum score (12 symptoms) of nervous exhaustion over trial period shown as means and standard error (SE), ITT-population (n = 154); ratings 0 = absent, 1 = mild, 2 = moderate, 3 = severe, max. score: 36; V1: Visit 1, V2: Visit 2, V3: Visit 3.

A post-hoc analysis showed that the symptom sum score decreased significantly in both groups over time. Additionally, 10 out of 12 characteristic symptoms abated to a larger extent in the Neurodoron® group when compared to placebo. Two of them, the symptoms irritability (p = 0.020) and nervousness (p = 0.045), were significantly reduced (fig. 3). Also, the change in the sum score of the Neurodoron® group (baseline minus visit 3 = 4.3 ± 0.6) was more pronounced than in the placebo group (2.8 ± 0.7), without reaching statistical significance (p = 0.091).

**Fig. 3.**
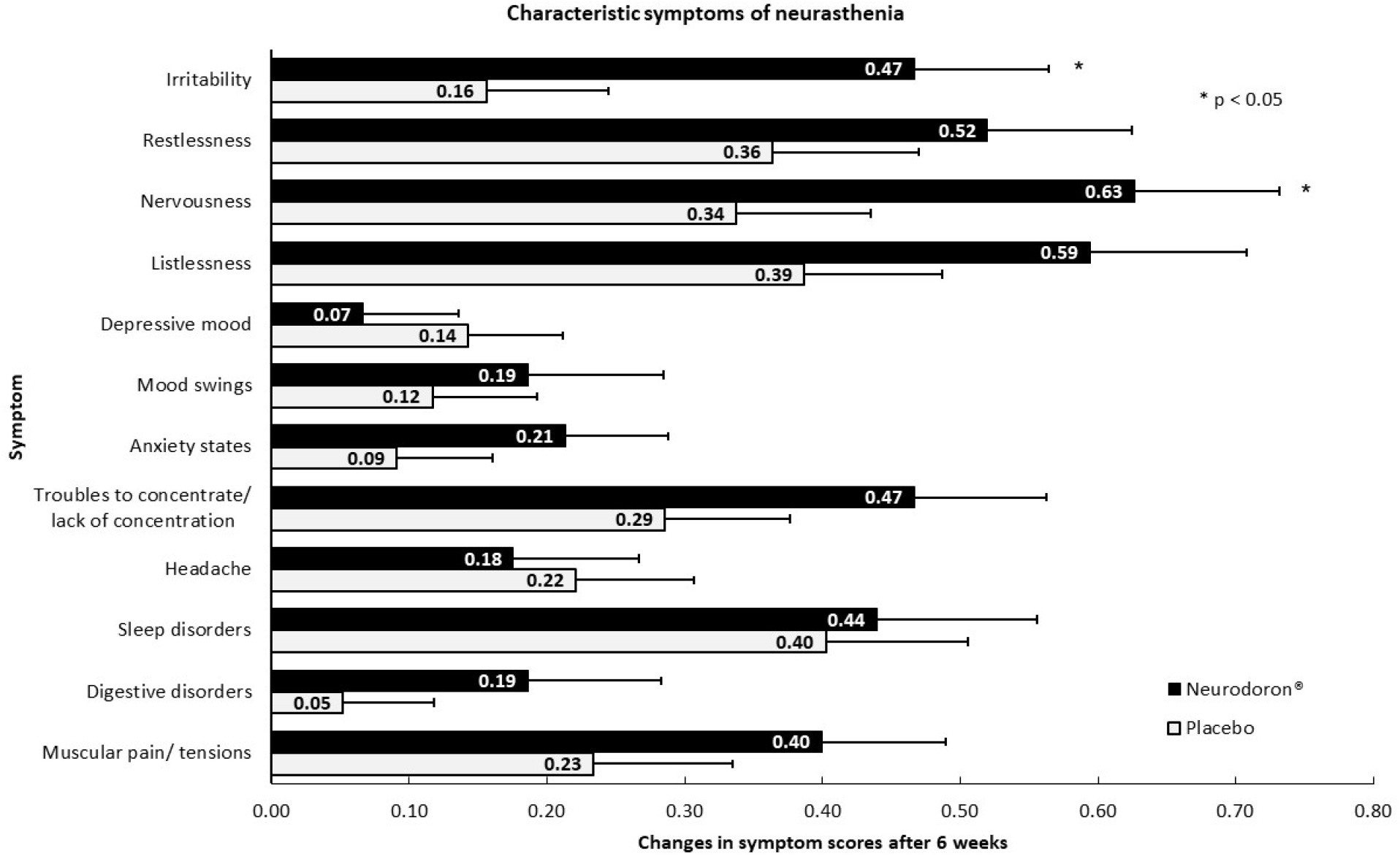
Post-hoc analysis, intra-individual differences (baseline minus visit 3) in the 12 characteristic symptoms of nervous exhaustion shown as mean and SE, ITT-population (n = 154); ratings per symptom: 0 = absent, 1 = mild, 2 = moderate, 3 = severe.

#### 4.2.2 Efficacy on perceived stress

At baseline perceived stress as measured with the PSQ was significantly higher in subjects of the Neurodoron® arm when compared to placebo in the a priori analysis. Post treatment no difference between groups was observed any more in the pre-specified analysis (p = 0.505, tab. 3, fig. 4).

**Fig. 4.**
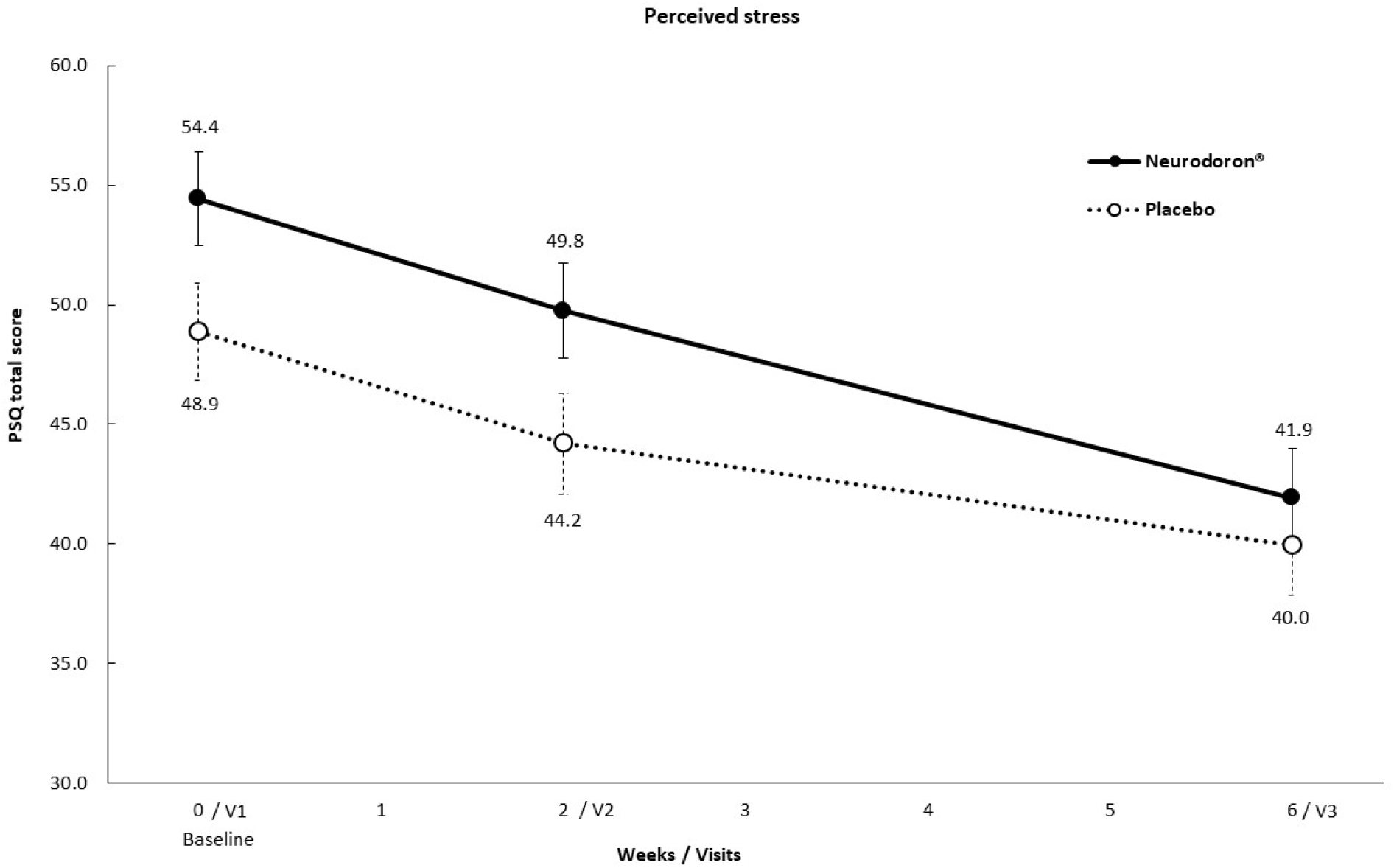
PSQ total score over treatment period of 6 weeks shown as means and SE, ITT-population (n = 154); total score between 0 to 100 (considering 4 subscales (worries, tension, joy, demands)). V1: Visit 1, V2: Visit 2, V3: Visit 3.

In both groups perceived stress decreased significantly over time on the overall score and on all subscales, as shown in the post-hoc analysis. A comparison of the PSQ subscales for pre-post treatment differences revealed a significantly greater increase of joy under Neurodoron® treatment (p = 0.037, fig. 5).

**Fig. 5.**
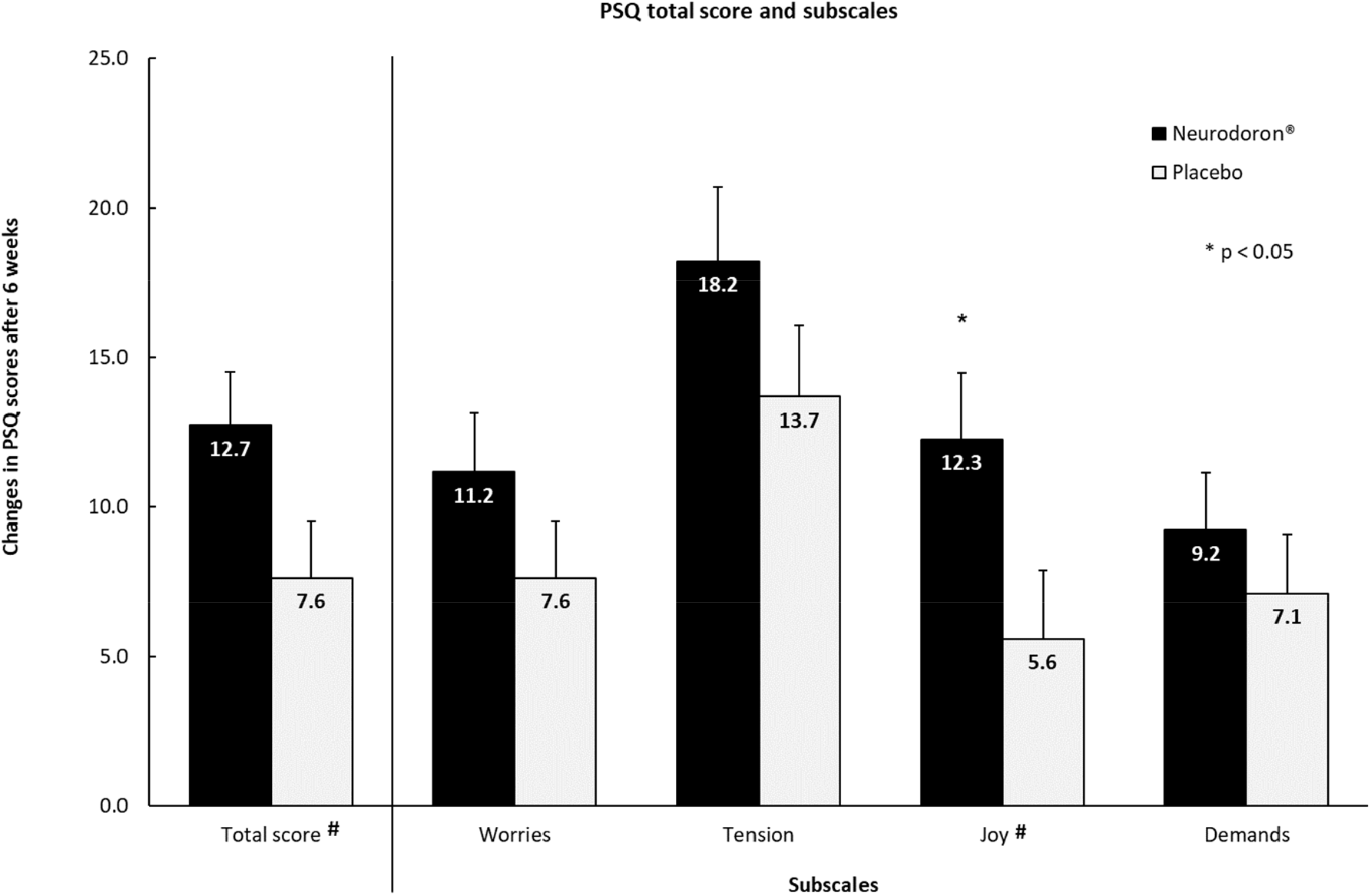
Post-hoc analysis, intra-individual differences (baseline minus visit 3) in the 4 PSQ subscales shown as mean and SE, ITT-population (n = 154); scores of subscales between 0 and 100. ^**#**^ ‘Joy’ values are inverted for calculation of total score and are shown inverted for better understanding, respectively.

#### 4.2.3 Health-related quality of life (SF-36)

Results of the SF-36 showed no significantly stronger improvement at visit 3, neither of the physical nor the mental sum scores for the Neurodoron® arm when compared to placebo (p = 0.773 (physical sum score), p = 0.510 (mental sum score), tab. 3, fig. 6a/b).

**Fig. 6a+b.**
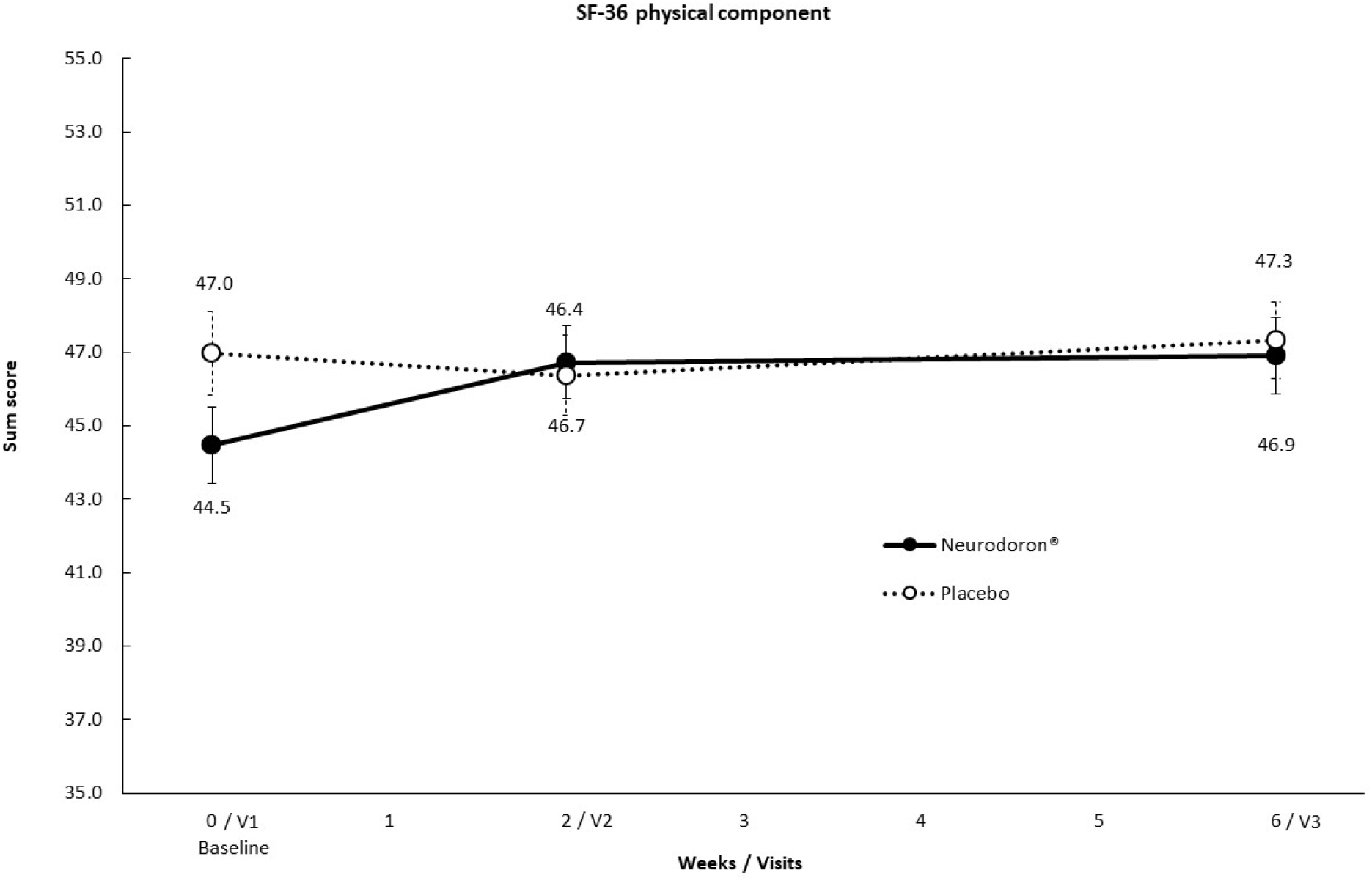
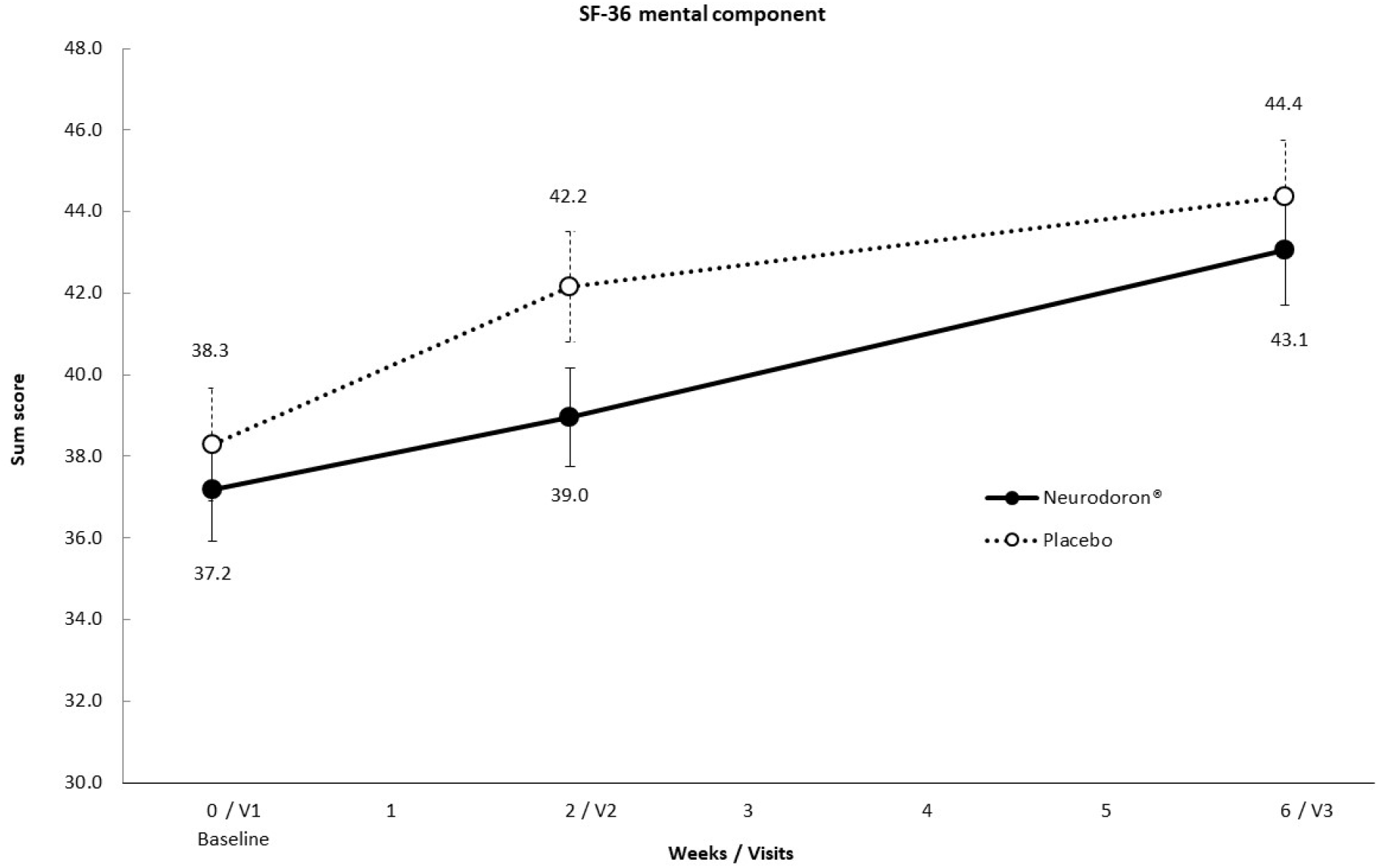
SF-36: Physical and mental sum scores (score range 0-100) over trial period shown as means and SE, ITT-population (n = 154). V1: Visit 1, V2: Visit 2, V3: Visit 3.

A post-hoc analysis showed that all subscales improved significantly over the treatment period, but the extent of improvement did not reach levels of statistical significance when comparing groups. Descriptively, patients treated with Neurodoron® had a greater improvement on the subscales social role functioning, vitality, mental health, general health perceptions, physical role functioning and physical functioning, while patients treated with placebo improved more regarding emotional role functioning and reported less bodily pain (fig. 7).

**Fig. 7.**
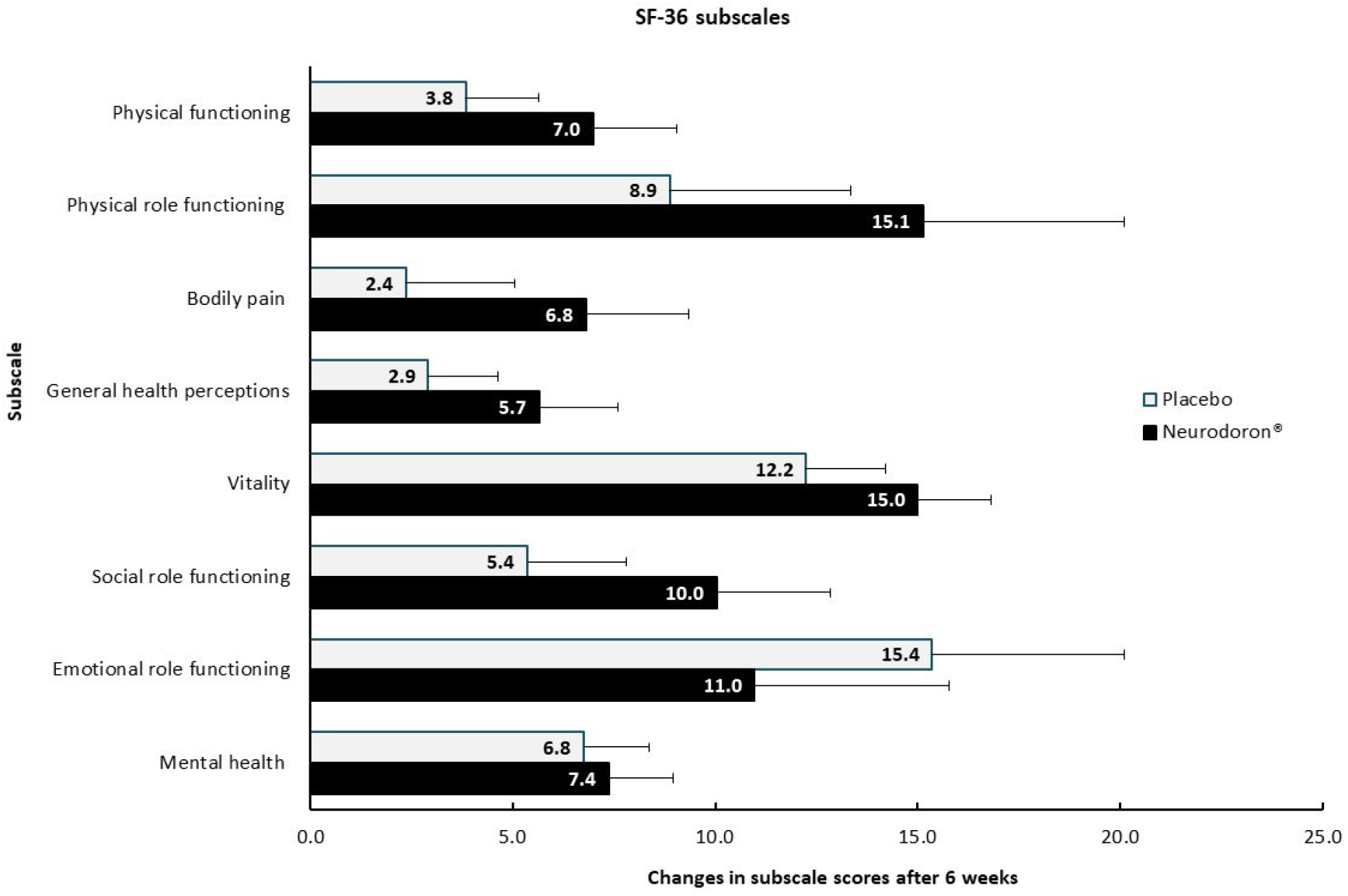
Post-hoc analysis, SF-36: Changes in subscale scores (score range 0-100) of 8 subscales over treatment period of 6 weeks shown as means and SE, ITT-population (n = 154).

The sensitivity analyses performed with the PP-population for all primary variables showed an equivalent outcome to the ITT analyses. The additionally performed supportive analyses with replaced missing values also confirmed the primary analysis.

### 4.3 Secondary efficacy and safety analysis

#### 4.3.1 Tedium Measure

In the a priori analysis, statistically significant differences between both groups were seen at baseline (p = 0.032) and visit 2 (p = 0.030) (fig. 8), with lower values for the placebo group, indicating a lower level of exhaustion. At visit 3 however, no statistical significance regarding group differences were seen.

**Fig. 8.**
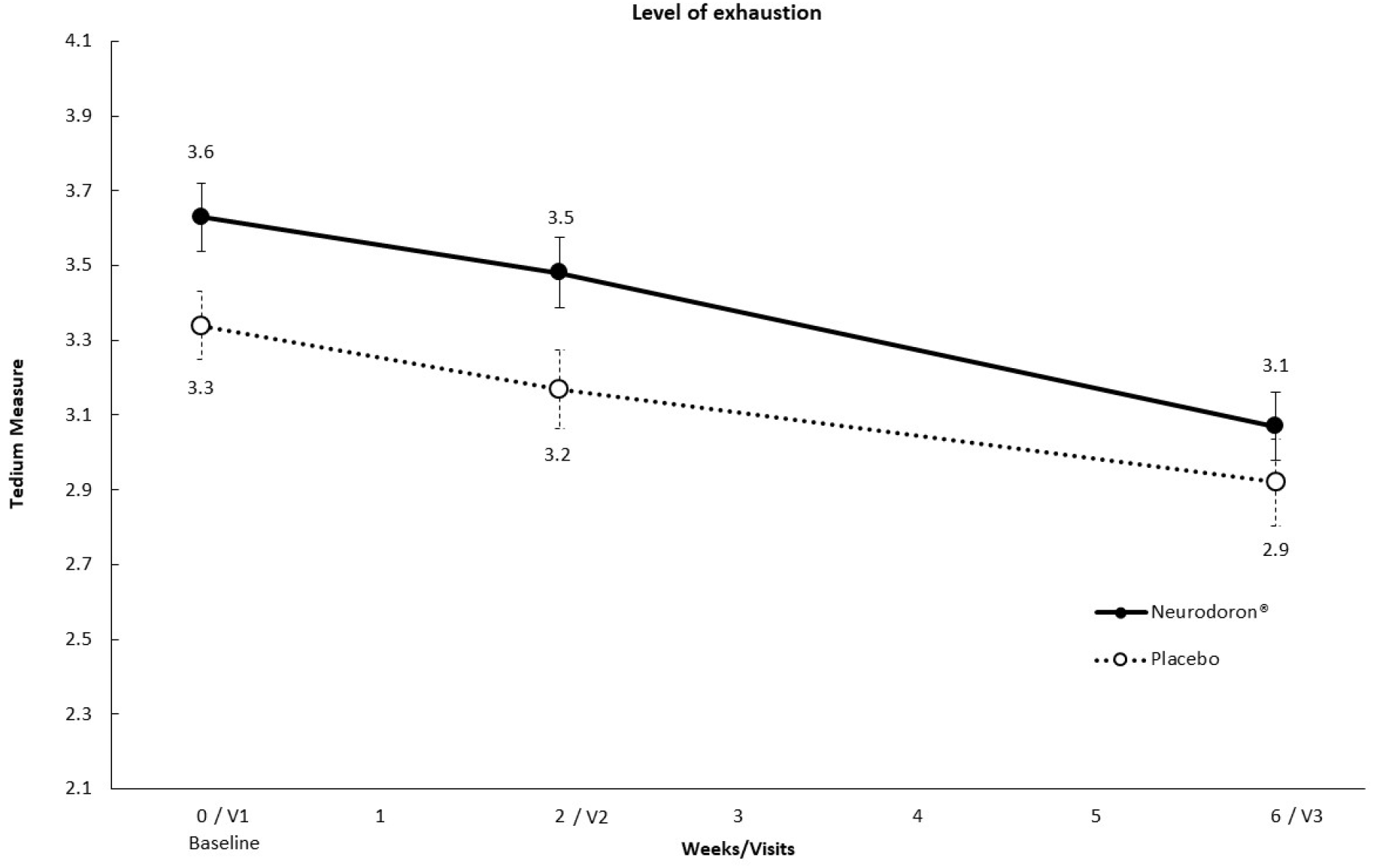
Tedium Measure (21 items) of level of exhaustion over trial period shown as means and SE, ITT-population (n= 154); Tedium Measure (min./max. score: 2.1/5.9). V1: Visit 1, V2: Visit 2, V3: Visit 3.

In a post-hoc analysis, results for the Tedium Measure showed a significant improvement for both groups over the course of the trial.

#### 4.3.2 Efficacy from patients and physicians’ point of view

51.3% of the patients evaluated the efficacy of Neurodoron® as good or very good. The efficacy of Neurodoron® was rated more frequently as good or very good by the patients than placebo (37.7%) without reaching statistical significance (fig. 9). The assessment of efficacy from the physicians’ perspective is very similar to that of the patients: 51.3% of physicians assessed Neurodoron® as good or very good, compared with 36.4% for placebo.

**Fig. 9.**
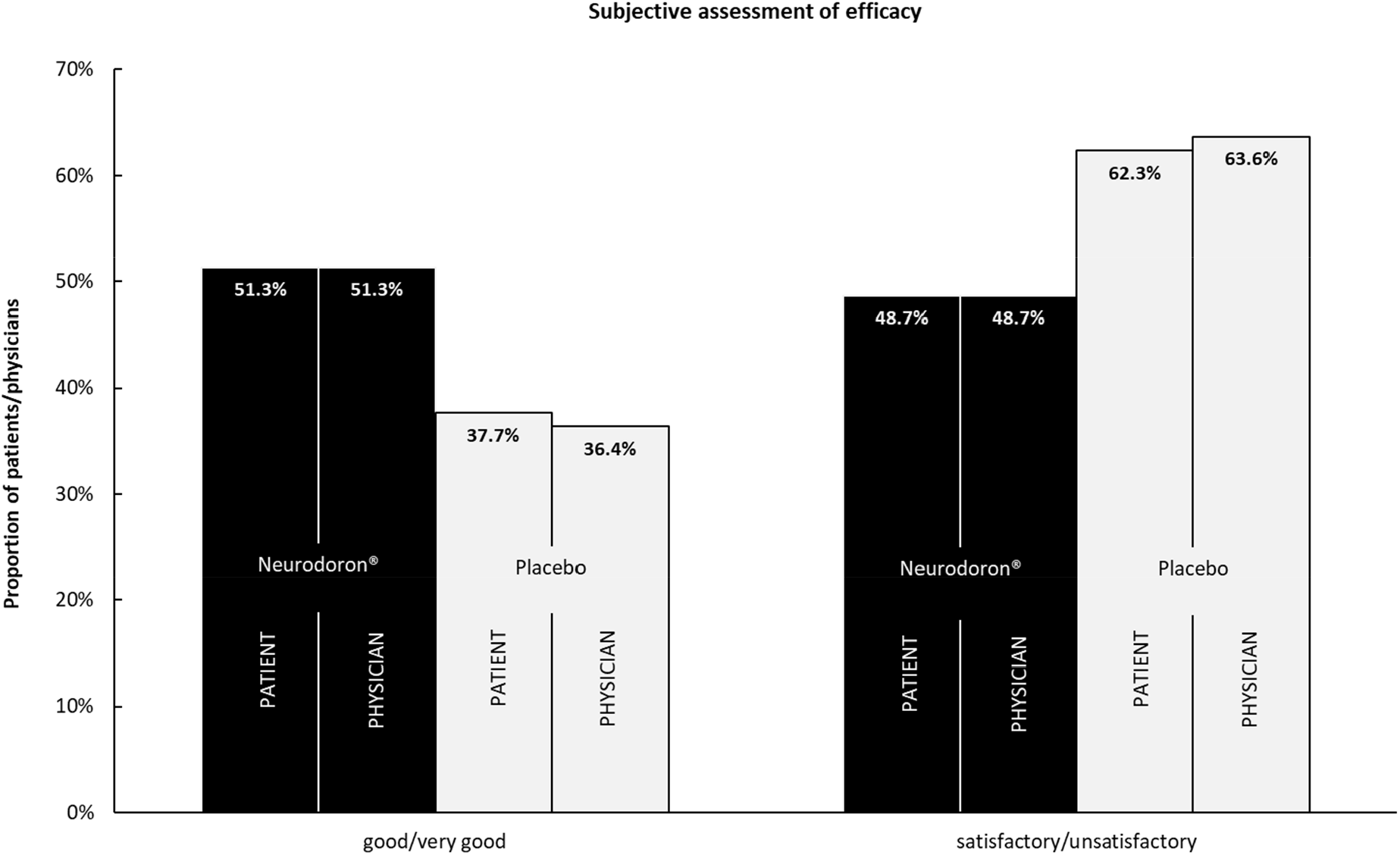
Subjective assessment of the efficacy of Neurodoron® (by patient and physician) at final visit after 6 weeks of treatment, ITT-population (n = 154); assessment options: very good, good, satisfactory, unsatisfactory.

#### 4.3.3 Subjective assessment of safety from patients and physicians’ point of view

96.1% of the patients evaluated the safety of Neurodoron® as good or very good vs. 97.4% in the placebo group (fig. 10). The physicians’ assessment was also in line with that of the patients: 97.3% of the physicians evaluated Neurodoron® as well or very well tolerated (97.4% for placebo treatment). 72.4% of the Neurodoron® patients would recommend the product (placebo group: 62.3%).

**Fig. 10.**
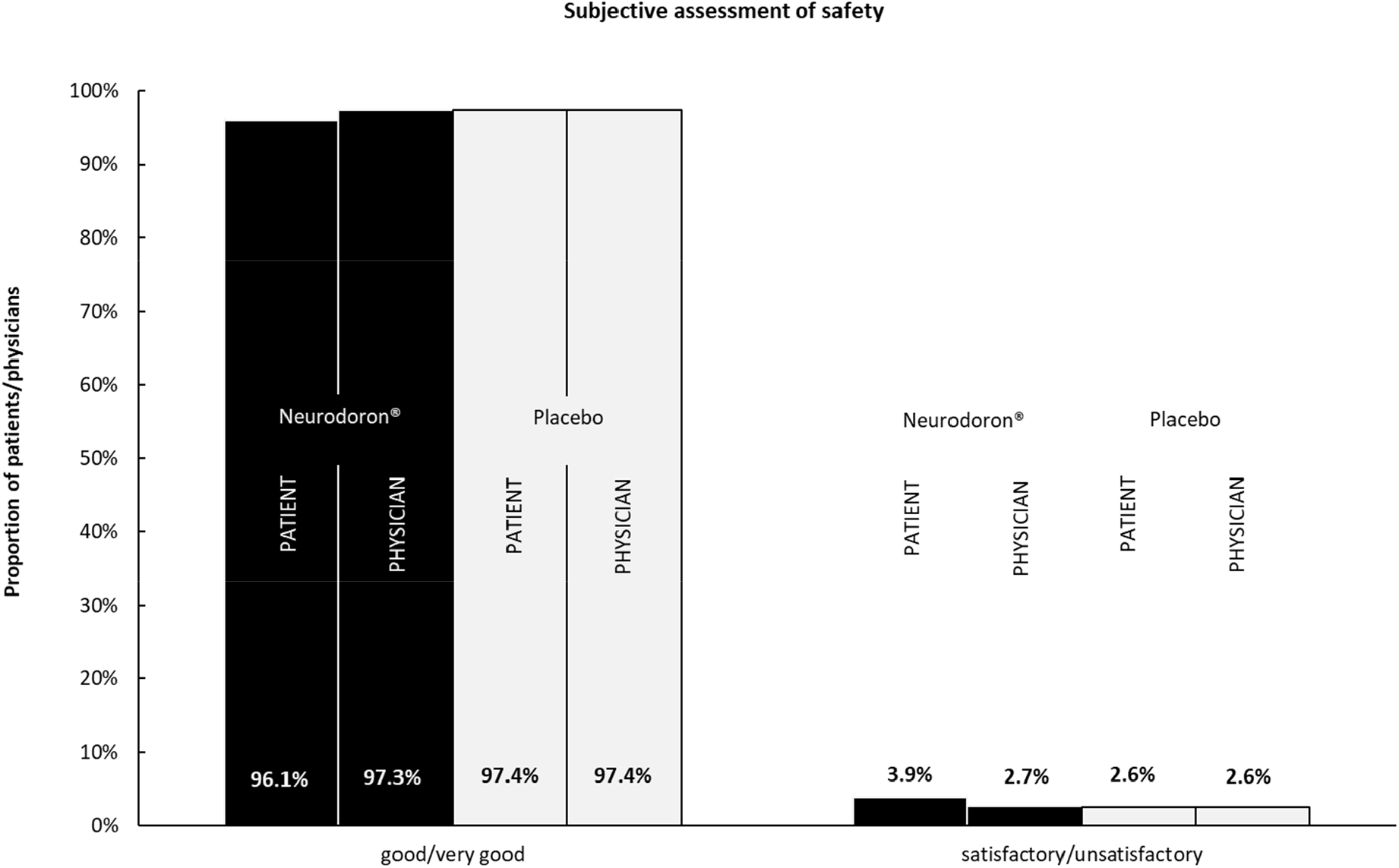
Subjective assessment of the safety of Neurodoron® (by patient and physician) at final visit after 6 weeks of treatment, ITT-population (n = 154); assessment options: very good, good, satisfactory, unsatisfactory.

#### 4.3.4 Safety evaluation of Neurodoron®

46 AEs (n = 32) were reported in the Neurodoron® group vs. 33 in the placebo group (n = 28). For both groups 6 AEs (Neurodoron®: n = 6, placebo: n = 5) were rated as causally related to the IMP. These AEs were all assessed as mild or moderate and mainly affected the gastrointestinal tract. One patient in the placebo group interrupted the IMP-intake, but there were no dropouts due to AEs or lack of efficacy. One serious adverse event occurred in the Neurodoron® group (postoperative joint dislocation). There was no causal relationship to the IMP. Overall, the most frequently reported AEs were diseases of the respiratory tract, chest and mediastinum (Neurodoron®: n = 13, placebo: n = 9), gastrointestinal tract (Neurodoron®: n = 4; placebo: n = 7), nervous system (Neurodoron®: n = 5, placebo: n = 4) and diseases of the skeletal muscles, connective tissue and bones (Neurodoron®: n = 6, placebo: n = 4). There were no signs for abnormalities in the blood tests.

## 5 Discussion

The first scientific mention of the diagnosis neurasthenia dates back to the 19^th^ century [1, 2]. Current prevalences for neurasthenia are between 1.3% and 10.8% [12]. One of the major reasons for neurasthenia is stress [1]. In the 2010s, adults report being more stressed than in the 1990s [37]. In parallel, mental illnesses are increasingly responsible for absenteeism from work; in Germany, for example, mental illnesses led to the highest number of sick days in 2020 [38]. This development increases the pressure on effective and safe treatment options.

Neurodoron® is an anthroposophic medicine for treatment of nervous exhaustion and metabolic weakness and has been on the market for over 50 years. In the authorization procedure, medical experience from long-standing use and specific characteristics were included, as for all medicinal products classified as phytotherapy, homeopathy, or anthroposophic medicine. Clinical data from NIS are available for Neurodoron®, the present RCT aims at confirming efficacy. While RCTs with their high internal validity are regarded as gold standard when it comes to proving efficacy of medicinal products, non-interventional trials are well perceived as supplementary sources of information, especially because they provide high external validity, i.e. their results can be generalized to all affected patients [39, 40]. A meta-analysis by Concato et al. even pointed out that the results of well-designed observational studies do not systematically overestimate the magnitude of treatment effects compared with those in RCTs on the same topic [41].

In the present RCT from 2011/2012, 204 patients were screened and 154 patients (77 in each group) were analyzed. Regarding baseline demographics, the two treatment groups differed only in terms of employment status and smoking behavior. Neither factor has yet been shown or is expected to be the sole cause of nervous exhaustion. Therefore, homogeneity of both groups at the beginning of treatment concerning demographics can be assumed. The proportion of women (73.4%) and the mean age of enrolled patients (53.1 years) was comparable to the two NIS conducted with Neurodoron® (physician-based NIS [24]: 78.0% and 50.3 years vs. 73.9% and 44.8 years in the pharmacy-based NIS [25]). Regarding symptom severity at baseline, patients of the Neurodoron® group showed a slightly more severe symptomatology of neurasthenia compared to the placebo group, with significant differences for the parameters perceived stress (PSQ) and Tedium Measure. This may have impacted the efficacy analysis.

The efficacy results of this double-blind RCT showed a significant improvement of neurasthenia after treatment of 6 weeks in both treatment groups for the investigated efficacy parameters. This is remarkable, as patients were suffering from persistent and distressing complaints of neurasthenia for more than 3 months prior to trial enrollment according to the ICD-10 criteria F48.0 [29]. However, the results did not achieve showing superiority of Neurodoron® over placebo in any of the pre-specified outcome measures. When looking at the course of the efficacy endpoints over the 6 weeks treatment time, Neurodoron® seems to be associated with a more pronounced improvement compared to placebo, without reaching significance in any of the a priori defined endpoints. Exemplary, 10 out of 12 characteristic symptoms of neurasthenia decreased more in the Neurodoron® group. Interestingly, a post-hoc analysis revealed some statistically significant group differences in favor of Neurodoron®, e.g. it showed that this anthroposophic medicinal product successfully reduced the characteristic stress symptoms nervousness and irritability. The intraindividual change after treatment with Neurodoron is similar for irritability (RCT: 0.5 scores, NIS: 0.6 scores) and nervousness (RCT: 0.6 scores, NIS: 0.5 scores). This indicates that the effect sizes between RCT and NIS are comparable, which is in line with Concato’s conclusion that effect sizes are not necessarily overestimated in NIS [41]. Furthermore, it improved the globally perceived stress in the subscale “joy”. It may well be that a larger sample size would have shown superiority of Neurodoron® over placebo in the pre-specified efficacy parameters. Sample size calculation of the RCT was based on the positive efficacy results of a physician based NIS performed with Neurodoron® in 2008/2009 [24]. The pharmacy-based NIS from 2014-2016 confirmed the good efficacy results of the physician-based NIS.

Three major differences between the two clinical study designs are likely to have contributed to the different findings. First of all, the NIS were not controlled. Therefore, the placebo effect is likely to reduce the magnitude of the Neurodoron® efficacy in the RCT. The placebo effect is particularly well documented for pain treatment [42-47]. In an inverse placebo RCT the placebo effect size was investigated. Patients who were told that they were being treated with a verum showed a decrease in pain perception, whereas patients who knew that they were receiving placebo had an unchanged pain perception 6 weeks after baseline as at baseline [48]. As neurasthenia has a strong mental component, it is likely that this indication is also prone for a strong placebo response. The effect size of placebo cannot be determined by the chosen design of the clinical trial. However, the results of the characteristic symptoms of neurasthenia observed in the RCT and the pharmacy-based NIS might be interpreted in this way. A second difference in the study design is the indication. Typical for an RCT, the inclusion diagnosis had clear and narrow criteria (F48.0 ICD-10) resulting in a more controlled setting. For the past physician-based NIS [24] as well as for the pharmacy-based NIS that was performed after this RCT [25], the inclusion indication was broader and referred to patient-reported stress-related disorders, according to the authorized indication. Therefore, patients may have presented to the NIS with both less severe as well as more severe symptoms of nervous fatigue. As Neurodoron® is an OTC product, it can be assumed that the patients of the pharmacy-based NIS best represent the affected patient population. Due to the rigorous patient selection in the RCT, it is possible that the treatment effect of Neurodoron® in the RCT differs from the effect seen in the two NIS. Lastly, the design of the NIS allowed a holistic treatment of this complex indication. From a CAM perspective, disease management pursues an overall therapeutic approach including recommendations concerning lifestyle changes to improve patient’s quality of life as a whole. In the RCT, patients were instructed to keep medication as for baseline, and they were prompted not to change their usual habits regarding daily activities such as sports, non-medicamentous therapies etc. As neurasthenia, or nervous exhaustion, is often due to mental complaints / chronic stress, a holistic approach that addresses the root of the complaints, is likely to show a stronger effect than treating the symptoms alone.

As shown above, the positive results of both NIS could be reproduced in the pre-specified primary efficacy evaluation of the RCT. However, as neurasthenia also improved in patients of the placebo group, the primary efficacy endpoint was not met. For the secondary efficacy parameters, the results are similar: The patients’ constitution significantly improved in both groups until treatment end, but no significant differences were found between groups. For patients in the Neurodoron® group, changes over the course of the trial indicate greater improvement in symptoms for all efficacy endpoints. For example, though not as convincing as efficacy ratings in the NIS, Neurodoron’s® subjective efficacy rating was more frequently considered by physicians and patients to be “good” or “very good” (both 51.3%) than placebo (37.7% and 36.4%).

Regarding safety, very few and non-severe side effects were reported for the entire trial. Overall, no differences were observed between Neurodoron® and placebo in the safety evaluation. The subjective safety rating of Neurodoron® was extraordinarily good with values between 96.1% (patients) and 97.4% (physicians) and assessment of AEs did not suggest any specific risks. These safety data confirm the available very good safety profile from the NIS and the continuous market monitoring with regard to product safety via the sponsor’s pharmacovigilance system.

## 6 Conclusions

This double-blind RCT adds information on an anthroposophic remedy and completes the overall picture consistently substantiating that Neurodoron® helps relieve stress related symptoms. This supports Neurodoron® being a good choice with regard to CAM approaches in patients with neurasthenia. Safety of Neurodoron® can be assessed as markedly good. RCTs with a comparable design and a higher number of patients may well clearly show that Neurodoron® is superior to placebo in the treatment of neurasthenia. Future clinical studies could strengthen outcomes by including biomarkers for a stress related dysregulation of the physiological system.

## 7 Statements

### 7.1 Ethics approval

This clinical trial has been registered on the “German Clinical Trials Register” (ID: DRKS00003261) and on EudraCT as 2010-024189-23. The trial protocol, informed consent document(s), and any other appropriate trial-related documents were reviewed and approved by an independent Ethics Committee and the relevant regulatory authorities.

### 7.2 Consent

All trial patients gave their written informed consent to participate in the clinical trial prior to any trial related procedures.

### 7.3 Acknowledgements

The authors would like to thank Bettina Bergtholdt, MD and director of emovis GmbH, a dedicated study site situated in Berlin, Germany, for the conduct of the RCT and Jörg Korb (Managing Director of ACM Allied Clinical Management GmbH) as well as Almut Günzel, Sandra Hildebrandt and Stefan Franke (former employees of ACM) for their excellent trial management and monitoring. The authors’ thanks also go to Hansjörg Wesp (MD, former employee of Weleda AG), Martin Schnelle (MD, Weleda AG) and Antje Gottberg (MD, former employee of CSG Clinische Studien Gesellschaft, IGES Group) for their contribution to the trial design and input for any medical matters. Also, we are grateful to Nicole Weber and Katja Schmidt ((former) employees of daacro GmbH & Co. KG) for their support in writing the manuscript. Special thanks go to all the anonymous patients for their valuable contribution to the trial. The trial was financed by Weleda AG.

### 7.4 Authorship confirmation statement

RH was responsible for conception and design of the trial and CS took care of data analyses and their correctness as well as data presentation. LS and JH drafted the manuscript and contributed substantially to the interpretation of the data. All authors reviewed the manuscript rigorously and critically and approved the final article.

### 7.5 Conflicts of interest

RH, CS and LS are employees of Weleda AG, Germany. JH is the CEO of daacro GmbH & Co. KG, a clinical research organization in Germany and declares having received an honorarium for writing the publication. No further interests have to be declared.

### 7.6 Funding information

The trial was financed by the Weleda AG, Arlesheim, Switzerland.

### 7.7 Data availability statement

The data that support the findings of this clinical trial are not publicly available due to containing information that could compromise the privacy of research participants. Disclosure of data would not be in accordance with the European General Data Protection Regulation.

## Supporting information

CONSORT Checklist

## Notes

### Clinical Trial

DRKS00003261

### Author Declarations

Ethical approval was granted by the Landesamt für Gesundheit und Soziales, Ethik-Kommission des Landes Berlin, Postfach 310929, 10639 Berlin, Germany

