## Supplemental Material I and II for "Neurodoron® in patients with neurasthenia – A randomized, double-blind, placebo-controlled clinical trial"

### I: Trial procedures and flow chart

Trial procedures:

| **Procedure** | **Trial Visit** | | | |
|---|---|---|---|---|
| | Screening week -4 to -1 | Baseline (Visit 1) week 0 | Visit 2 week 2 (± 3 days) | Visit 3* week 6 (± 5 days) |
| Patient information and patient consent | X | | | |
| Demographic und anthropometric data | X | | | |
| Anamnesis (exclusion of organic reasons, major depression, major anxiety disorder) | X | | | |
| Physical examination | | X | | X |
| ECG | X | | | |
| Vital signs | X | X | X | X |
| Blood sampling | X | | | X |
| Diagnosis (criteria of neurasthenia) | X | | | |
| Pretreatment | X | | | |
| Concomitant medication, concomitant disease | X | X | X | X |
| Inclusion criteria | | X | | |
| Exclusion criteria | | X | | |
| Assessment of characteristic symptoms (neurasthenia) | | X | X | X |
| Self-assessment scales (PSQ, SF-36, Tedium Measure) | | X | X | X |
| Randomization | | X | | |
| IMP: dispense and return | | X | X | X |
| IMP treatment phase | | \|--------------------------------\| | | |
| Assessment of compliance | | | X | X |
| Adverse events | | | X | X |
| Efficacy and safety evaluation (patient, investigator) | | | | X |

* Visit 3 is also to be done in case of premature discontinuation, if possible.

Flow chart:

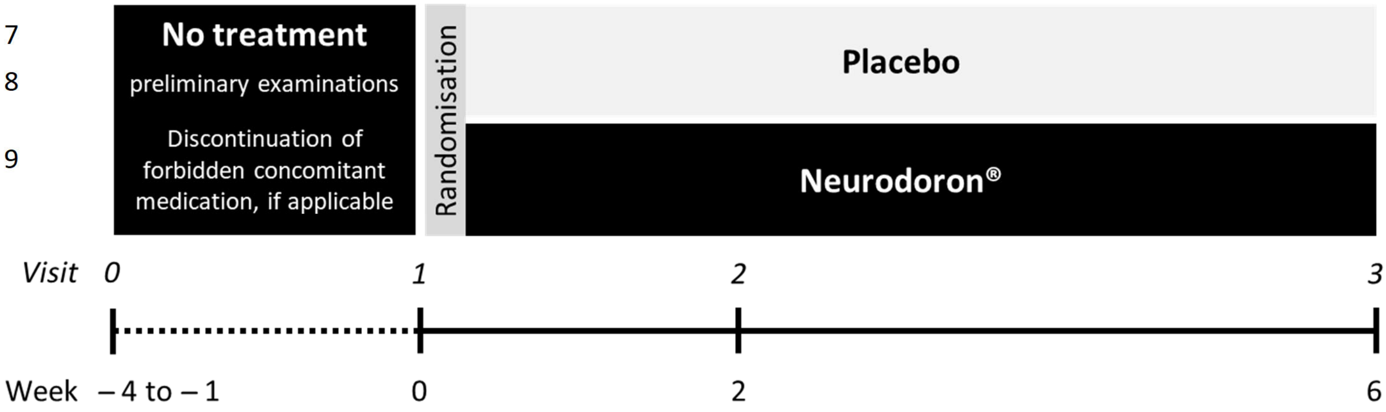
No treatment
preliminary examinations
Discontinuation of forbidden concomitant medication, if applicable
Randomisation
Placebo
Neurodoron®
Visit 0
1
2
3
Week – 4 to – 1
0
2
6

### II: Inclusion/exclusion criteria

#### Inclusion criteria:

1. Presence of Signed Informed Consent Form
2. Age ≥18 years
3. Nervous exhaustion (neurasthenia) characterized by:
   - A. *either 1. or 2.*
     1. persistent and distressing complaints of increased fatigue after minimal mental effort (e.g., after accomplishing or attempting to accomplish everyday tasks that do not require unusual mental effort)
     2. persistent and distressing complaints of exhaustion and bodily weakness after minimal effort

   *and*

   - B. at least one of the following symptoms: acute or chronic feelings of muscular aches and pains, dizziness, tension headaches, sleep disturbances, inability to relax, irritability.
   - C. The individual concerned is not able to recover from A.1 or A.2 within a normal period of rest, relaxation, or distraction
   - D. The duration of the complaints is at least three months.

#### Exclusion criteria

1. Concurrent or not at least 4 weeks past participation in other clinical trials involving the use of an investigational medicinal product or non-drug interventions for the treatment of nervous exhaustion
2. Known hypersensitivity to wheat starch
3. Organic reason for complaints (e.g. known heart, kidney, liver disease, poorly controlled infection diabetes mellitus, possibly relevant iron deficiency anemia [Hb-value <12 g/dL], hypothyroid metabolic state)
4. Neuropsychiatric disease (z. B. psychosis, schizophrenia, dementia or other diseases that cause symptoms of exhaustion)
5. brain organic disorders (e.g. postencephalitic syndrome, organic psychosyndrome following traumatic brain injury)
6. Suspicion of major depression, defined by BDI-II (Beck Depression Inventory) ≥ 29
7. Suspicion of major panic disorders or generalized anxiety syndrome, defined by Generalized Anxiety Disorder 7-item (GAD-7) score ≥ 16

8. Treatment with forbidden medication, e.g., other medications for stress/burnout/exhaustion, intake of antidepressants within the past 4 weeks, intake of benzodiazepines within the past 4 weeks, initiation of treatment with oral β-blockers within the past 4 weeks
9. Known abuse or addiction to drugs, alcohol, or prescription drugs
10. Placement in an institution due to official or court order
11. Pregnancy, lactation
